# Clinical Evaluation of Ultrasonicated ADSCs in Atrophic Non-Union of Long Bones

**DOI:** 10.1101/2025.07.17.25331624

**Authors:** Himanshu Bansal, Alnkrita Bansal, Irfan Khan, Kristia Chrysostomou, Anupama Bansal, Jerry Leon, Mustafa al Maini, Sanjay Singh

**Author notes:** Corresponding author Dr. Himanshu Bansal, MD, Consultant Regenerative medicine, Revita Lifesciences, Rudrapur, India-263153.

## Abstract

**Background:** Non-union is a common clinical complication that causes serious limb debilities and eventually leads to permanent limb impairments if not treated on time. This proof-of-concept study was undertaken to evaluate the osteogenic potential of autologous adipose derived stromal cells (ADSCs) in long bone non-union.

**Methods:** Seven patients with long-standing atrophic non-union of long bones were treated with percutaneous implantation of a minimum of 100 million ADSC and platelet-rich plasma containing a minimum of 10 billion platelets into the non-union site of long bones under fluoroscopic guidance. Besides radiological imaging for evidence of consolidation, patients were also followed up for clinical parameters, standard lower extremity functional scale (LEFS) and SF12 scores. The patients were followed for a minimum of one year, & Intervention was considered a failure if no evidence of healing was observed up to 6 months post-procedure

**Results:** We observed union in 6 of 7 (86%) patients within 4 months after the procedure. The first evidence of healing was visible within 8 weeks after treatment in five (71%) patients.

**Conclusions:** The study demonstrated the efficacy of ADSCs as a better alternative to bone marrow aspirate concentrate (BMAC) in treating long bone non-union.

**Level of Evidence:** Level 3

**Trial Registration:** This study has been registered in the US Clinical Trial Registry (U.S. National Library of Medicine) with Trial registration no. NCT04340284. Titled as “Adipose Derived Stromal Vascular Fraction (SVF) Application in treatment of Long Bone nonunion” https://clinicaltrials.gov/ct2/show/NCT04340284?term=NCT04340284&draw=2&rank=1

Date of registration 09/04/2020

## 1. Introduction

Non-union is an unfortunate complication in fracture management and is considered established when at least nine months have passed since the fracture without any sign of union [1]. Based on radiological appearance, non-union fractures are described as hypertrophic, atrophic, and oligotrophic, among which atrophic is considered most complicated as it is inert and avascular with no biological potential to repair compared to other types of non-union [2]. Bone grafting with or without disconnecting the fibrous union has been the gold standard [2, 3] It has several disadvantages, such as increased morbidity, pain, paresthesia, blood loss, risk of re-fracture, infection at the donor site, a limited amount of cancellous bone, and devascularization of the fracture fragments [3–5]. Allografts and bone morphogenic protein, besides high cost and lower engraftment capacity, also inherit the possibility of rejection and transmission of diseases [6–8].

The osteo-inductive properties of mesenchymal stem cells (MSCs) in bone formation as a minimally invasive treatment of non-union have gained a lot of interest in the recent past [9, 10]. The MSCs residing in the local environment of the fracture have the potential to re-ossify fractures, as these cells are in a latent state and require growth-stimulating conditions to regulate the healing microenvironment of the tissue [11]. Bone marrow aspirate concentrate (BMAC) has captured the attention for the treatment of non-union fractures because of the presence of a heterogenous cocktail of MSCs, growth factors, and various types of progenitor cells [12–14]. However, bone marrow harvesting procedures are painful and, at the same time, yield a low MSC count [15]. Recently, adipose-derived stromal cells (ADSCs) have emerged as an alternative source of MSC in a single point of care procedure [15–18]. Besides the advantage of an easy harvest of adipose tissue under local anaesthesia, adipose tissue contains 500–2500 times more MSCs compared to an equivalent volume of bone marrow [19]. Interestingly, the number of mesenchymal stem cells does not decrease with age in adipose tissue, and MSCs isolated from this tissue are considered genetically more stable with higher differentiation capacity [19].

The effect of ADSCs has been extensively studied in animal models with critical-size segmental defects, and the results show that it significantly enhances the rate of new bone formation both at the centre and the periphery of the defect [20, 21]. Although the earliest clinical application of autologous ADSC, also known as stromal vascular fraction (SVF), in non-union, was reported in 2004 [22] Only a few case reports and clinical studies with ADSC application in non-union have been published to date [23–27].

The present study aims to evaluate the practicality of minimally invasive percutaneous injection with autologous adipose-derived stem cells (ADSCs) and explore their osteoinductive efficacy in the management of non-union. To the best of our knowledge, there are no reasonably large case studies reported in long bones’ atrophic nonunion; hence, this work will provide insight into an adipose tissue-based resolution for non-union fractures.

## 2. Materials and Methods

### 2.1 Eligibility and Patient recruitment

This study was ethically approved by the Institutional Committee for Stem Cell Research and Therapy at Anupam Hospital, Uttarakhand, India. The study adhered to the Consolidated Standards of Reporting Trials (CONSORT) guidelines, and informed consent was obtained from all participants before their inclusion. A total of 7 patients with long bone fractures who failed to unite even after 9 months of standard treatment were considered enrolled in this study to evaluate the safety and efficacy of fluoroscopy-guided percutaneous implantation of ADSC at the site of fracture as an outpatient procedure. This trial was registered in Clinicaltrials.gov-NCT04340284; Date of registration 09/04/2020; Clinicaltrials.gov under URL: https://clinicaltrials.gov/ct2/show/NCT04340284).

### 2.2 Patient recruitment for the study

A total of 18 patient records from individuals who underwent ADSC implantation at Anupam Hospital, Rudrapur, between 2012 and 2018 were reviewed for eligibility. Seven patients were excluded for not meeting the inclusion criteria, primarily due to active infection or a fracture gap exceeding 5 mm (Table 2.1). In addition, follow-up data were unavailable for four patients, who were also excluded. Consequently, only seven patients were included in the final analysis (Fig. 1). A total of 18 patient records from individuals who underwent ADSC implantation at Anupam Hospital, Rudrapur, between 2012 and 2018 were reviewed for eligibility (Fig.1).

**Table 2.1:**
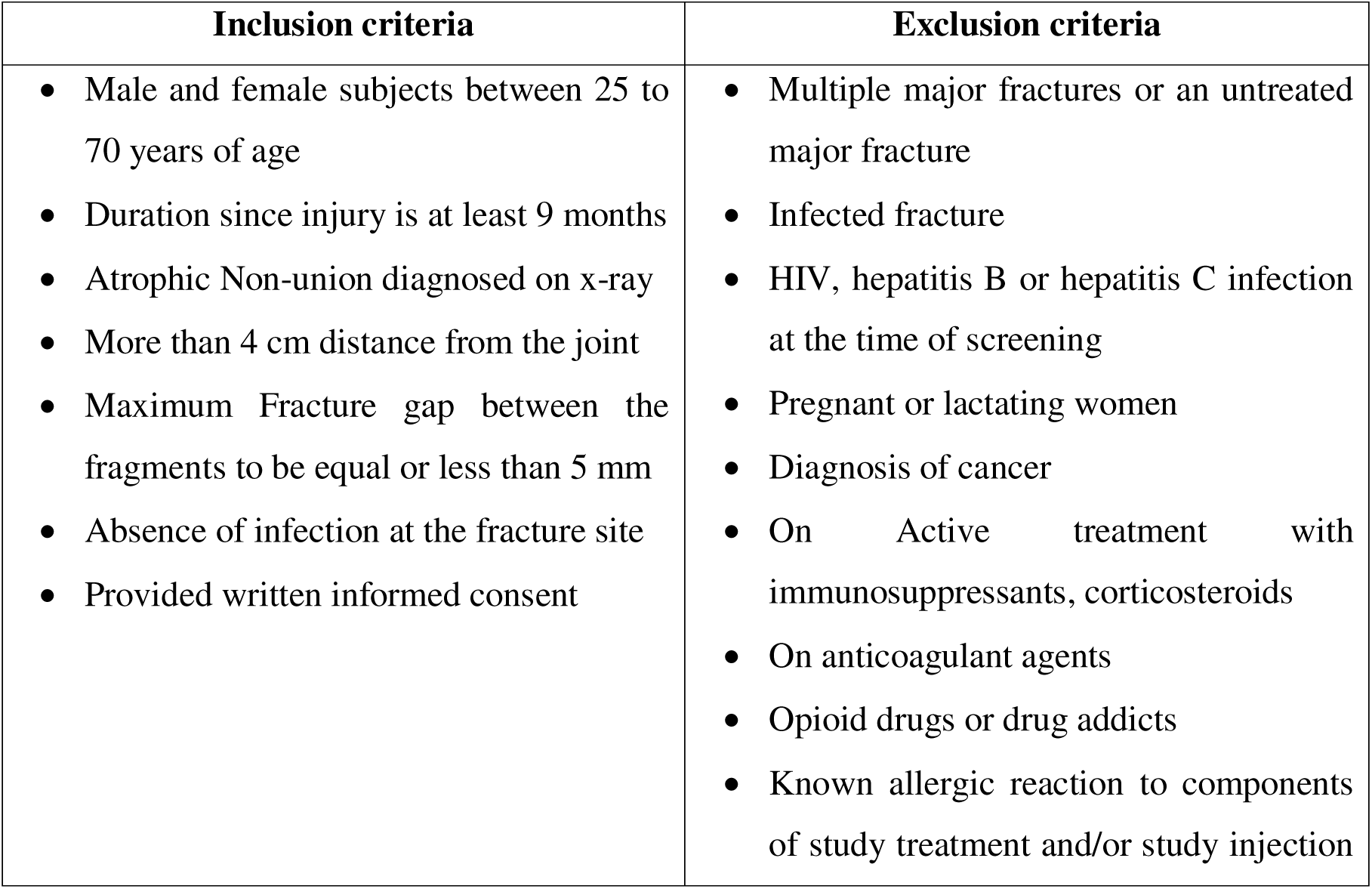
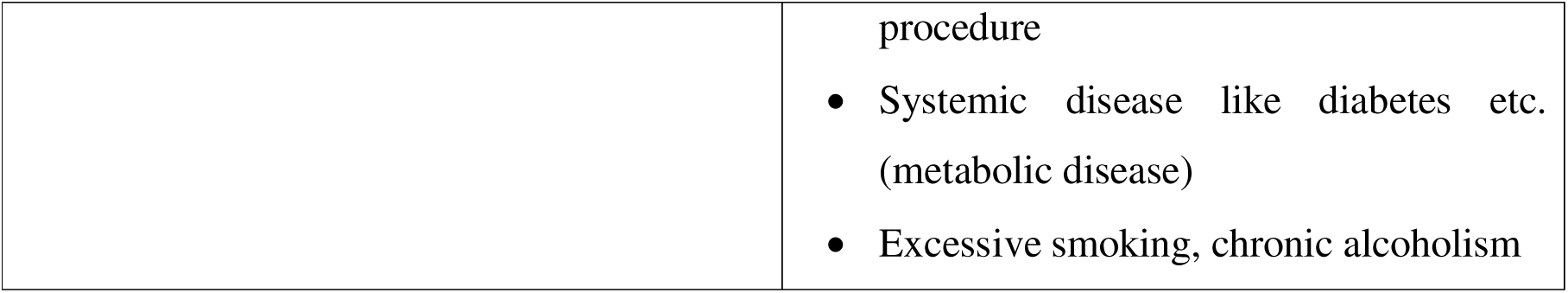
Eligibility criteria for enrolment into the study.

| Inclusion criteria | Exclusion criteria |
| --- | --- |
| • Male and female subjects between 25 to 70 years of age<br>• Duration since injury is at least 9 months<br>• Atrophic Non-union diagnosed on x-ray<br>• More than 4 cm distance from the joint<br>• Maximum Fracture gap between the fragments to be equal or less than 5 mm<br>• Absence of infection at the fracture site<br>• Provided written informed consent | • Multiple major fractures or an untreated major fracture<br>• Infected fracture<br>• HIV, hepatitis B or hepatitis C infection at the time of screening<br>• Pregnant or lactating women<br>• Diagnosis of cancer<br>• On Active treatment with immunosuppressants, corticosteroids<br>• On anticoagulant agents<br>• Opioid drugs or drug addicts<br>• Known allergic reaction to components of study treatment and/or study injection |
|  | procedure<br>• Systemic disease like diabetes etc. (metabolic disease)<br>• Excessive smoking, chronic alcoholism |

**Fig. 2.1.**
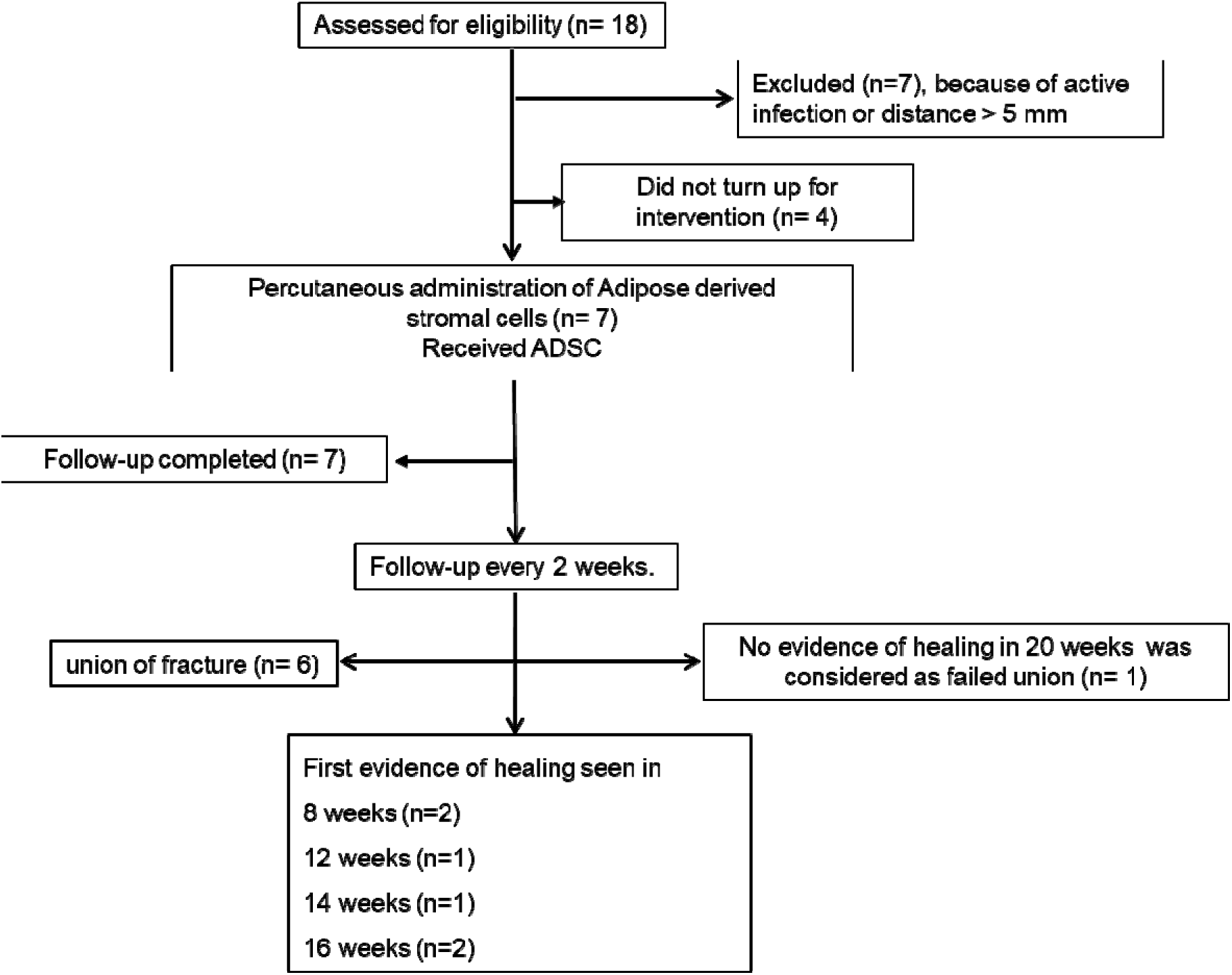
Flowchart of the clinical trial: Screening, assessment, treatment allocation and follow-up of patients with OA. (n = number of patients).

### 2.3 Criteria for non-union and assessment for infection

The indication for this procedure was established atrophic non-union with a maximum gap between the fragments to be ≤ 5 mm and duration of not more than one year between the occurrence of fracture and this procedure. The other main criteria were the absence of infection at the fracture site. Non-union was considered established when: I) At least 9 months have passed since the injury, and II) the fracture did not show any visible signs of healing in the previous 3 months [4]. Only atrophic non-unions were considered for enrollment. Screening tests were conducted to investigate for signs of active local or systemic infection. The fracture site was also subjected to hot fomentation for 7 days to flare up any latent infection, if any. Patients were enrolled in the study only if they met the eligibility criteria (Table 1). Plain x-ray anteroposterior and lateral views were taken every fort-night for radiological assessment, as it is still considered the gold standard to diagnose and follow-up fracture non-unions [28].

### 2.4 Isolation of adipose-derived stromal cells

In all the cases, adipose tissue was harvested under local anaesthesia with mild sedation of midazolam (1mg/kg slow intravenous). Approximately 60 cc of lipoaspirate was extracted from the patient’s abdominal fat using an aspiration cannula of 3 mm with prior administration of an equal amount of Klein tumescent solution. ADSCs were harvested from adipose tissue non-enzymatically using the sonication principle [29]. The lipoaspirates were transferred to 50 mL centrifuge tubes. The tubes were kept in an ultrasonic cavitation machine (Revita Lifesciences, Rudrapur, India) and homogenised under sterile conditions at 40 MHz for 10 minutes. To obtain the ADSCs, the sonicated homogenised mixture was centrifuged at 400g for 10 minutes.

The pellet was then resuspended in 5 mL saline and passed through a 100 µm mesh filter (Thermo Fisher Scientific, USA), then centrifuged again at 400g for 10 min to obtain a washed and cleaned ADSC pellet. The obtained pellet was resuspended in 1 mL of normal saline for each administration (Fig. 2.2). The cell number, viability, and purity of freshly isolated patient ADSCs were phenotypically characterised following reference guidelines [30, 31].

The total cell count and viability were determined using the automated MUSE© Cell Analyser (Merck Millipore, USA). Flow cytometric analysis was conducted to assess identity markers (CD90, CD73, and CD105) and purity markers (CD34 and HLA-DR) with the BD FACS Calibur™ system (BD Biosciences, USA).

**Fig. 2.2.**
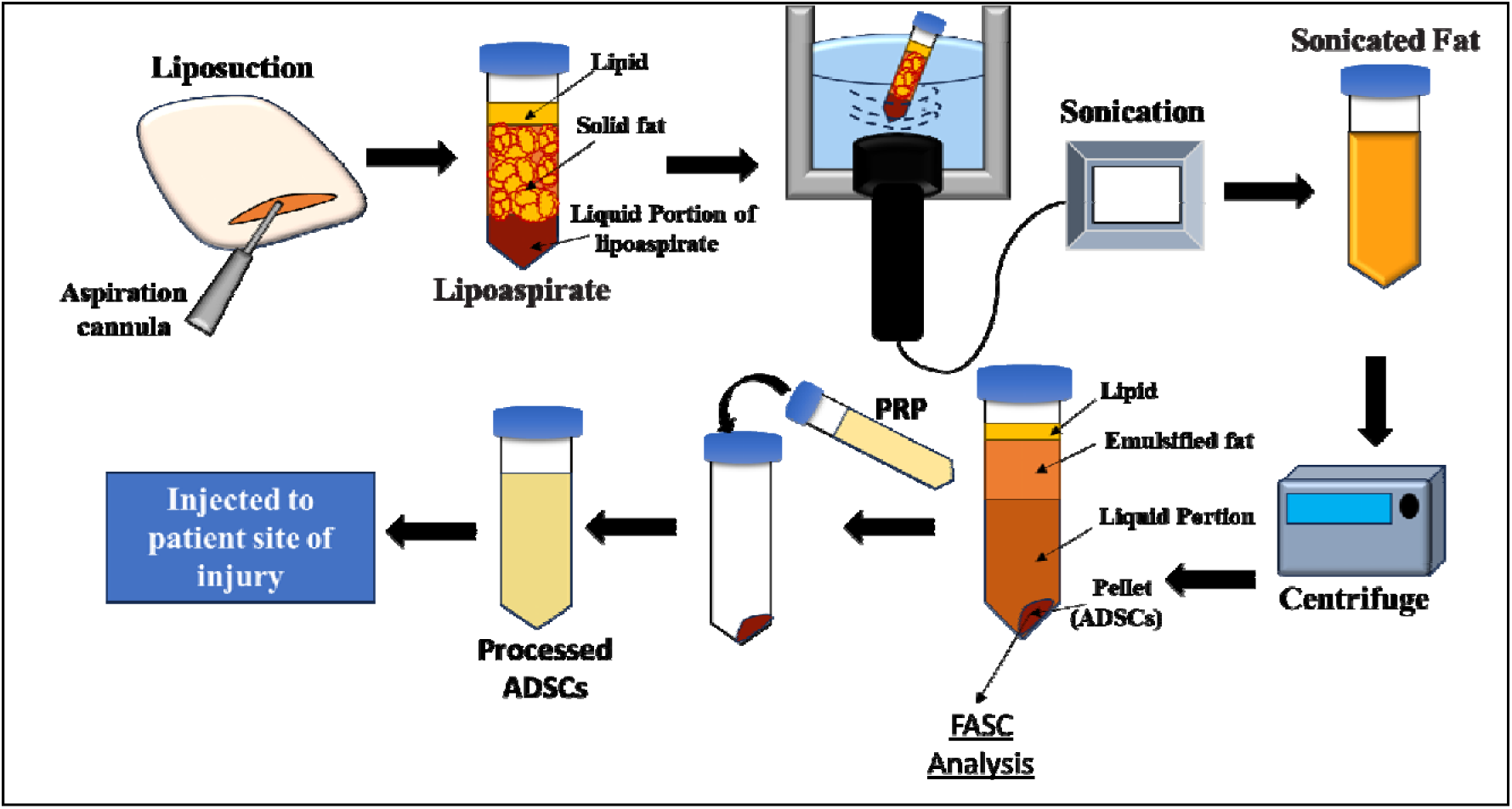
Schematic representation of the isolation and processing of ADSC before injecting to the patient.

### 2.5 Platelet rich plasma (PRP) preparation

Platelet-rich plasma (PRP) was derived from the peripheral blood of the same patient. Briefly, a 60 mL blood sample was drawn in 10 % ACD solution and centrifuged at 600xg for 10 minutes before collecting the plasma fraction. The obtained plasma fraction was centrifuged at 4000xg for 15 minutes. The supernatant (platelet-poor plasma) was then carefully removed, leaving 7 mL of PRP. Platelet-poor plasma was also filtered with a 1 µm special flush-back filter to trap remaining platelets and mixed with the PRP [32]. In line with established guidelines [33], the prepared PRP was characterised for quality and cellular composition, including platelet, leukocyte, and red blood cell (RBC) counts, using the automated haematology analyser Sysmex XN-10 (Sysmex, Kobe, Japan).

### 2.6 Preparation of the fracture site and fluoroscopic localisation, and Percutaneous injection

The fracture site was cleaned with alcohol, betadine, and draped. Local anaesthesia was applied around the site. With a mobile C-Arm machine (Siemens), anterior-posterior and lateral views were used to identify the site of injections. 8 mL of solution having ADSCs and PRP was injected using (18G needles; Bectan Dickinson and Company, NJ, USA) into the non-union site and around the bones without removing the fibrous tissue. Compression strapping was done to prevent discharge or hematoma.

### 2.7 Assessments of Patient Recovery

Healing at the injury site was evaluated clinically and radiologically at every 2 weeks until 12 months post-treatment. Anteroposterior radiographs with the joint fully extended were obtained as standard procedure. Two independent, blind experts (orthopaedic surgeon and radiologist) reviewed the x-ray images to determine whether there was any evidence of persistent non-union or signs of union of the fracture. The radiological progression was determined as per the scoring method[10], and the scores were used to quantify non-union and healing. Patient-reported outcomes were also assessed using the Short Form 12 (SF-12) and the Lower Extremity Functional Scale (LEFS).

Tolerability and safety were assessed by monthly evaluation of haematological and biochemical parameters, as well as by detailed clinical examination of any local or systemic adverse events if reported by the patient. Adverse events were categorised as isolated, intermittent, or continuous and depending on their occurrences as mild, moderate, or severe based on the interference with the patient’s daily activities. A possible causal relationship with the supplement in terms of Definite/Possible/Probable/Non-Assessable/None was also assessed.

### 2.8 Statistical analysis

The statistical significance was assessed by Student’s t test using 2-tailed Mann–Whitney U test. Data are expressed as medians (range). *P-*values less than 0.05 were considered statistically significant.

## 3. Results

### 3.1 Demographics

The age of the patients ranged from 25 to 58 (median 48) years. Five of the 7 patients were male. The clinical characteristics of these patients have been described in (Table 3.1)

**Table 3.1.** Summary of patient characteristics.

| S. No | Characteristics | Values |
| --- | --- | --- |
| 1. | Age (years) | 25-58 years |
| 2. | Sex | Male -5; Female – 2 |
| 3. | Mean (SD) | 45 (12) years |
| 4. | Median | 48 years |

All patients underwent initial treatment with standard closed/open reduction and internal/external fixation at the time of primary management (Table 3.2). All conservative attempts, like functional bracing, partial weight bearing, and dynamization, had already been unsuccessfully attempted in all patients to facilitate union before embarking on percutaneous ADSCs injection.

**Table 3.2.** Clinical characteristics of patients at the time of enrollment.

| **Patient No** | **Type of fracture** | **Initial treatment** | **Duration of nonunion (Before beginning ADSC treatment)** |
| --- | --- | --- | --- |
| P1 | Tibia Open II | Debridement and external fixator were later treated at the 3rd month with bone grafting, locked IM nailing | 14 months |
| P2 | Tibia Open IIIa | Debridement and external fixator. External fixation was removed in patient P2 after the wound healed at 4 months and the fracture was stabilised with an above-knee POP cast for 3 months, followed by a Patellar tendon bearing walking POP cast for another 3 months. | 10 months |
| P3* | Tibia Closed | Closed interlocked nailing | 9 months 10 days |
| P4 | Tibia Closed | Closed interlocked nailing | 9 months |
| P5* | Femur Closed | Interlocked nail presented with loosening of screw and persistent instability at fracture | 9 months |
| P6 | Femur Closed | Locked plating | 12 months |
| P7 | Operated femur; infected | External fixator after removal of locked IM nail. The infection was successfully treated before their treatment. | 16months |

\* Female patients

**Table 3.3.**
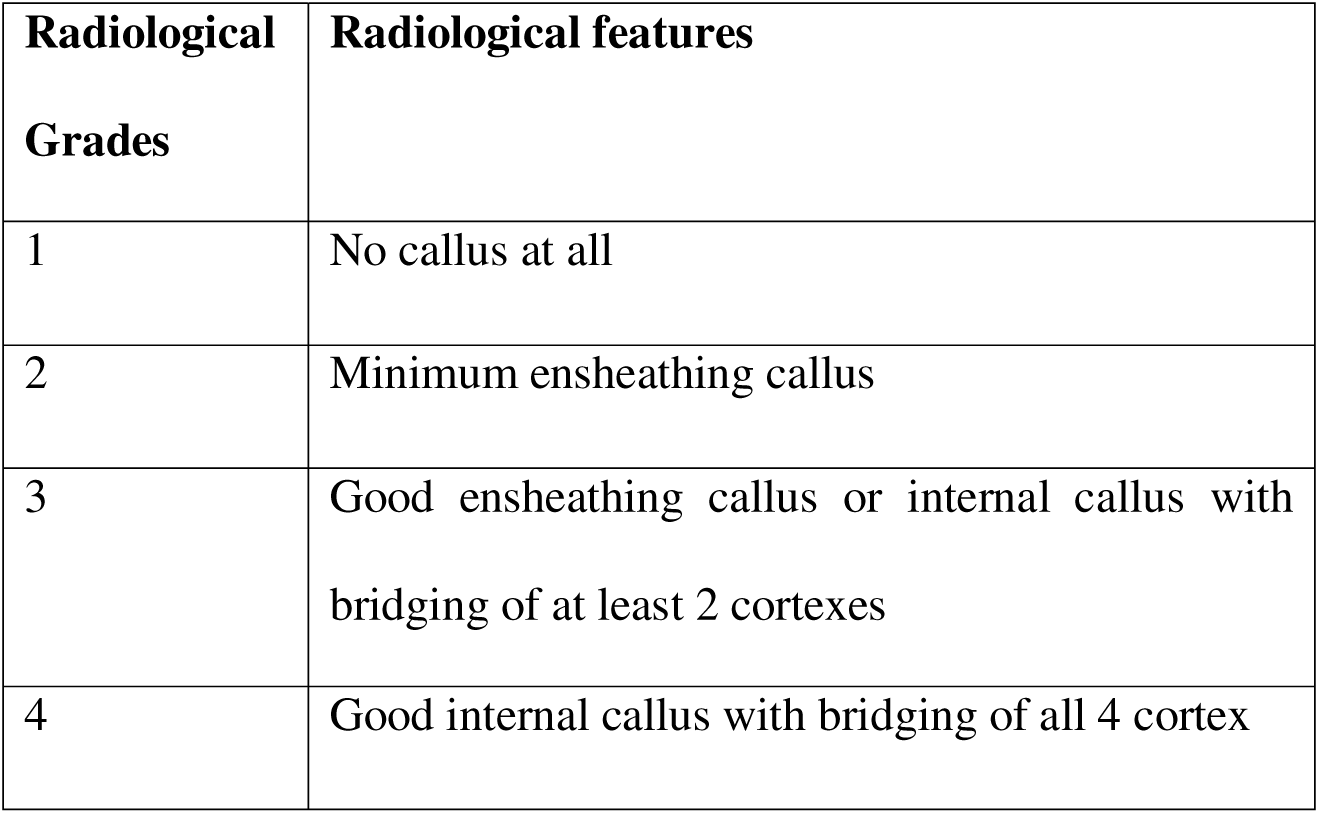
Radiological grading for fracture healing.

### 3.2 Biological characteristics of ADSCs concentrate-PRP injections

Using our isolation protocol, we successfully obtained approximately 1–2 million adipose-derived stem cells (ADSCs) per cubic centimetre (CC) (Table.4). The freshly isolated uncultured ADSCs phenotype was characterised for multiple positive and negative cell surface markers using FACS analysis. The ADSCs isolated from the patient tissue shows CD29^+^ (90-97%), CD73^+^ (92-94%), CD90^+^ (90-98%), and CD105^+^ (83-88%) as a positive marker for ADSCs (Fig. 3.1) and CD14^-^, CD31^-^ and HLA-DR^-^ as a negative immunophenotype markers (Fig. 3.1).

**Fig. 3.1.**
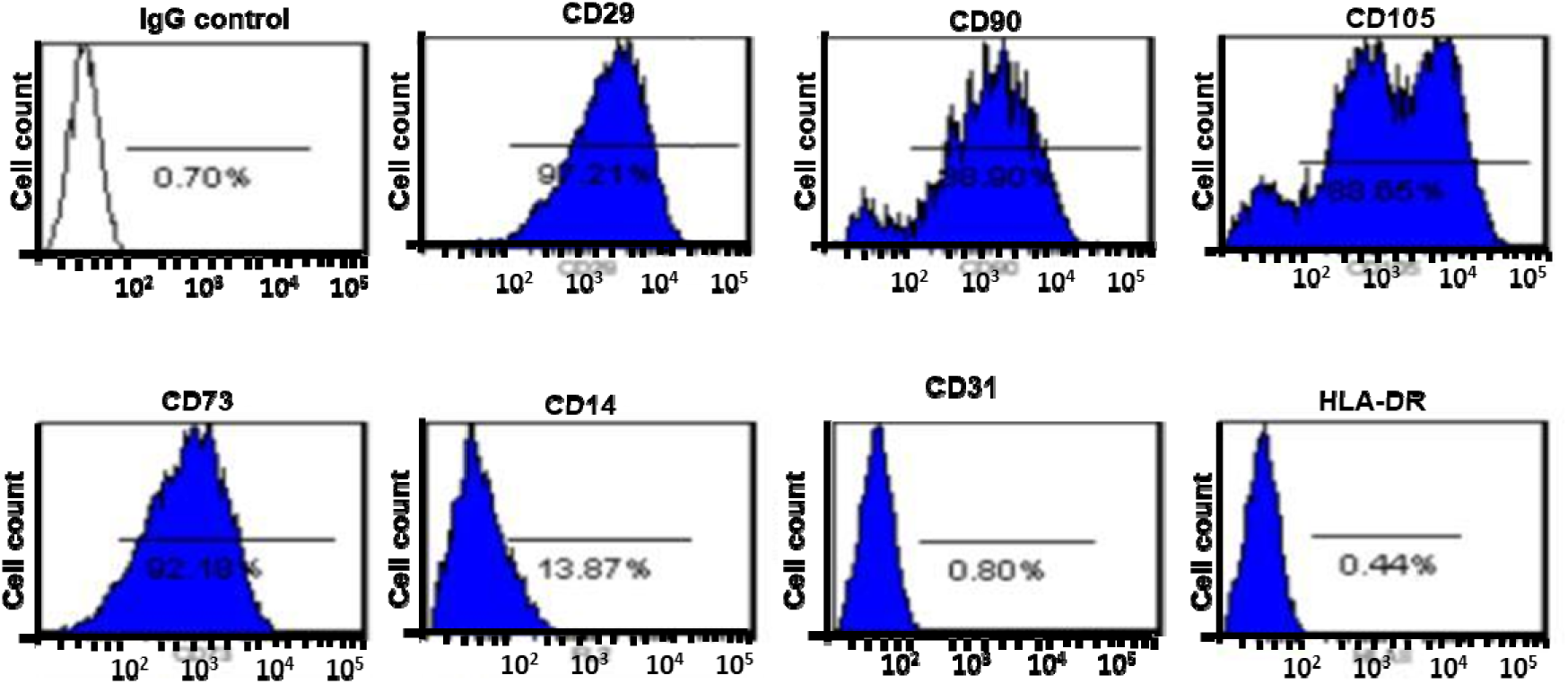
Flow cytometry analysis of ADSCs; Immunophenotype expression of CD29, CD73, CD90 and CD105 were expressed at very high levels, while CD14, CD31 31 and HLA-DR at very low levels.

**Table 3.4.** Individual patient counts of administered ADSCs and platelets.

| No. of Patients | ADSC harvested<br>(counts per one mL in millions) | Absolute number of ADSC Injected (in millions) | Platelet Counts per mL | Absolute number of platelets injected |
| --- | --- | --- | --- | --- |
| P1 | 102 | 102 | 14.4 x10<sup>8</sup> | 100.8 x10<sup>8</sup> |
| P2 | 108 | 108 | 13.2 x10<sup>8</sup> | 92.4 x10<sup>8</sup> |
| P3 | 102 | 102 | $14.6 \times 10^8$ | $102.2 \times 10^8$ |
| P4 | 120 | 120 | $13.8 \times 10^8$ | $96.6 \times 10^8$ |
| P5 | 110 | 110 | $16.1 \times 10^8$ | $112.7 \times 10^8$ |
| P6 | 106 | 106 | $13.4 \times 10^8$ | $93.8 \times 10^8$ |
| P7 | 118 | 118 | $14.2 \times 10^8$ | $99.4 \times 10^8$ |

Each patient received a minimum of 100 million stromal cells, with a total ADSC count ranging between 102-120 million cells, along with at least 10 billion platelets (Table 3.4). A total volume of 8 mL, consisting of ADSC concentrate combined with platelet-rich plasma (PRP), was administered percutaneously using an 18-G needle. The injection was delivered at four circumferential points around the non-union site under fluoroscopic guidance to ensure precise placement. The time from initial injury to non-union or delayed union and ADSC treatment ranged from 9 months to 16 months. After ADSC implantation, six of the seven fractures united within 4 months of injection. A single injection was found to be sufficient in all cases.

The patients showed evidence of healing as early as two months, with full consolidation on average after 6-8 weeks after the first documented sign (Table 3.5).

Follow-ups were performed every 2 weeks for a minimum of 12 months following the procedure. Significant improvements were noted in LEFS (54.6-86.7; *P* <0.02), as well as Short Form 12 (SF12) health survey score (27.2-48.4; P<0.05).

### 3.3 Case description

Two patients with open tibial fractures (P1 and P2) were treated initially with debridement and external fixation. Patient P1 was later treated at the 3^rd^ month with bone grafting, locked IM nailing before presenting as non-union at 14 months. This patient showed first evidence of healing at 16 weeks and full consolidation in the next 12 weeks (Table 3.5)

External fixation was removed in patient P2 after the wound healed at 4 months and the fracture was stabilised with an above-knee POP cast for 3 months, followed by a patellar tendon bearing walking POP cast for another 3 months. However, at 10 months fracture remained unstable, although the wound had completely healed but with poor skin condition. There was an unsuccessful healing of the fracture, which required an Illizarov fixation, bone grafting and muscle flap coverage. The reason for failure seems probably a severe open injury needing extensive wound care for 4 months. Poor vascularity at the fracture site also contributed to failure.

**Table 5.** Case description of the study.

| **Case No.** | **Duration of nonunion before beginning ADSC treatment** | **Time to evidence of healing Grade 2** | **Further progression of healing to Grade 3** | **Full consolidation Grade 4** |
| --- | --- | --- | --- | --- |
| 1 | 14 months | 16 weeks | 24 weeks | 28 weeks |
| 2 | 10 months | None | - | None |
| 3 | 9 months 10 days | 16 weeks | 22 weeks | 26 weeks |
| 4 | 9 months | 12 weeks | 18 weeks | 22 weeks |
| 5 | 9 months | 8 weeks | 16 weeks | 22 weeks |
| 6 | 12 months | 8 weeks | 14 weeks | 22 weeks |
| 7 | 16 months | 14weeks | 22 weeks | 38weeks |

All time frame is post-ADSC implantation

Two patients with closed-shaft tibia fractures (P3 and P4) were dynamized at 2 months after primary closed interlock nailing before presenting as non-union at 9 months. Both these patients showed full consolidation in 26 and 22 weeks after ADSCs implantation.

Patients 5 initially treated with interlocked nailing presented with loosening of screw and persistent instability at fracture site by 9 months. Post ADSCs implantation, the first evidence of callus was seen in 8 weeks (grade 2 union), which matured to consolidation in the next 14 weeks (Table 3.5)

Patient P6 with supracondylar femoral fracture earlier treated with plates and screws (Fig. 3.2 a-b) however, even after 12 months this patient presented with non-union and implant failure (Fig. 3.2 c). Post implantation of ADSCs showed the first evidence of healing by 8 weeks (Fig. 3.2 d) and consolidation following another 6 and 8weeks (Fig. 3.2 e and f).

**Fig. 3.2.**
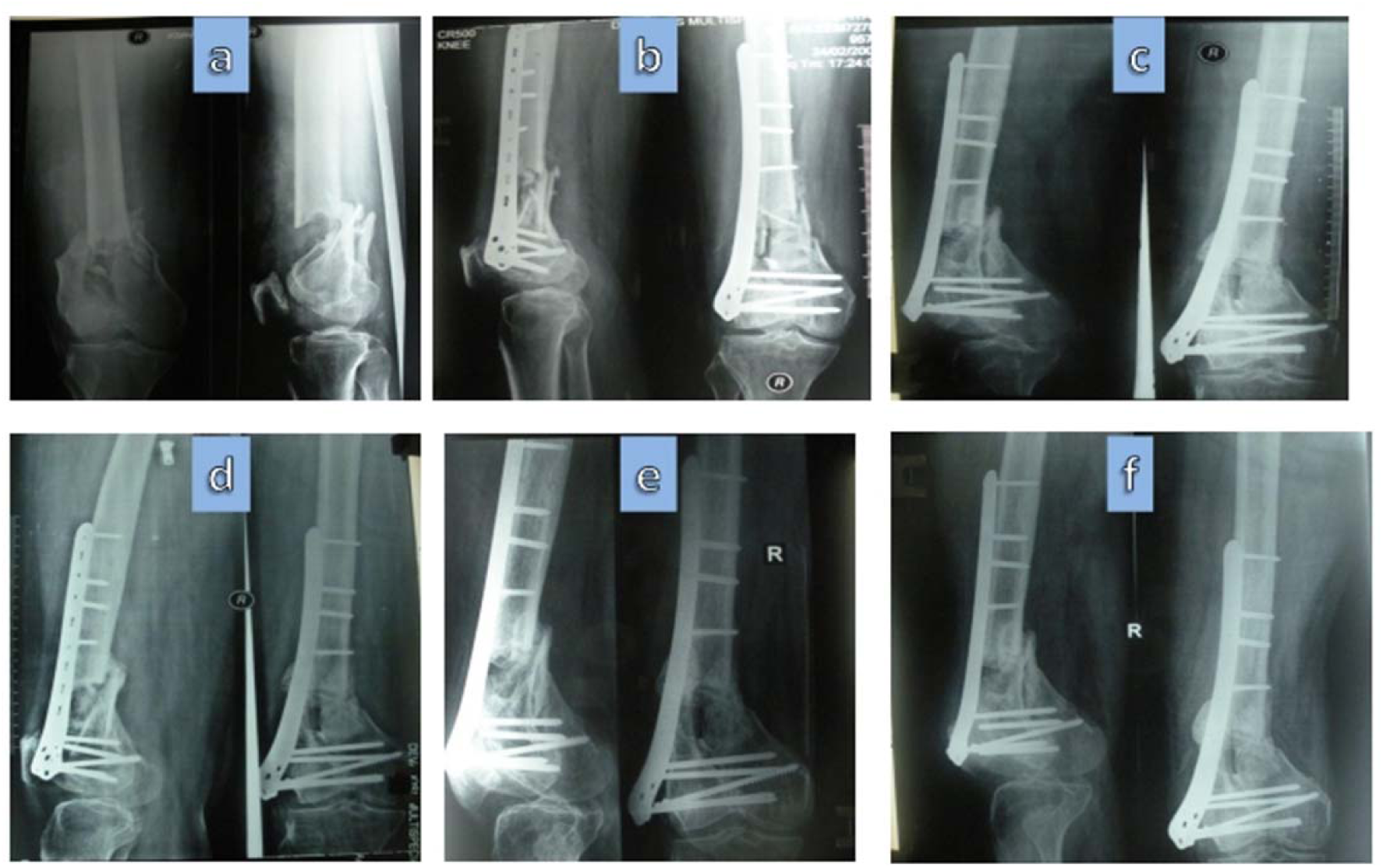
Serial anteroposterior and lateral plain x-rays showing healing of non-union in Patient P6 **(a)** fracture in supracondylar femur **(b)** well fixed fracture supracondylar femur with plates and screws **(c)** nonunion at baseline at 12 months post injury showing implant failure and non-union **(d)** post ADSC procedure 8 weeks showing beginning of the union **(e)** post ADSC procedure 14 weeks showing consolidation **(f)** post ADSC procedure 22 weeks showing mature consolidation.

Patient P7 suffered a post-operative infection following fixation of the femur fracture (Fig. 3.3 a). it led to the removal of nail and external fixator application (Fig.3.3 b) and was being treated for infection, which completely subsided only after 16 months (Fig. 3.3 c). An open procedure with revision was still not considered safe owing to the recurrence of the infection, so percutaneous ADSC implantation was performed. The first evidence of callus was noted by 3 months, and good consolidation by 28 weeks. The first evidence of healing was seen in the patient at 14 weeks (Fig. 3.3 d), and a good consolidation in the next 24 weeks (Fig. 3.3 e and f). This patient is archetypal of a successful outcome in the worst-case scenario

**Fig. 3.3.**
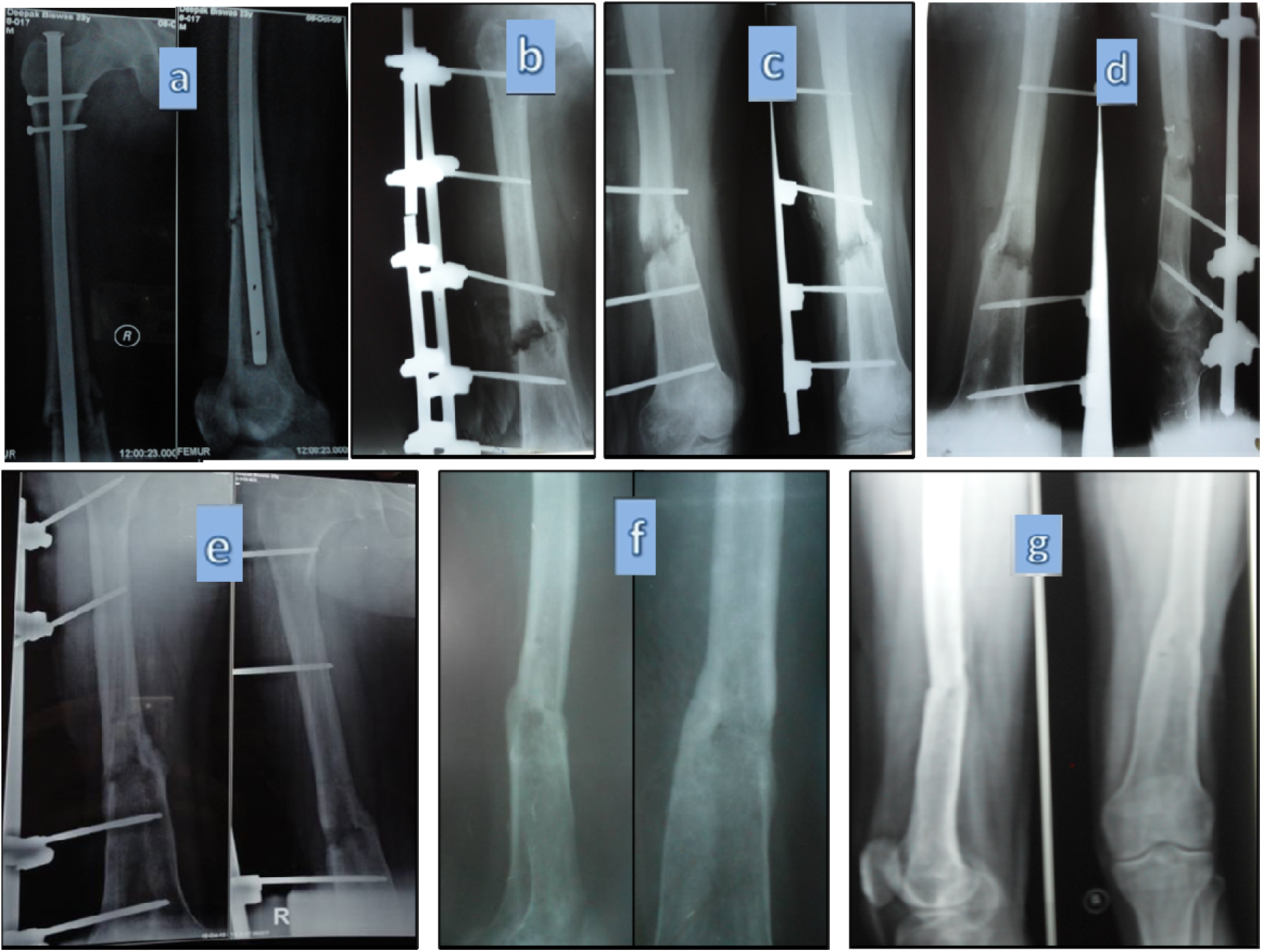
Serial anteroposterior and lateral plain x-rays showing healing of non-union in - Patient P7 **(a)** infected fracture femur with intramedullary nail in situ and **(b)** external fixation after IM nail removal 20 weeks post injury **(c)** baseline image at 16 months post injury after eradicating all possible injection **(d)** 14 weeks post procedure image showing initiation of healing **(e)** 18 weeks marginal improvement in healing without change in grade **(f)** 22 weeks healing reached grade 3 and **(g)** 38 weeks post ADSC healing reached grade 4.

## 4. Discussion

Autologous stem cell-based procedures have revolutionised a single-step surgical process for skeletal tissue regeneration [34]. Their direct application in the vicinity of the fracture helps in the differentiation and proliferation of osteoblast cells for new bone formation [35]. Mesenchymal stem cells (MSCs) have paracrine effects, which inhibit pro-inflammatory pathways, and promote tissue regeneration by releasing anabolic cytokines and differentiating into connective tissue cells [36]. Moreover, the biological mediators released by MSCs promote a regenerative microenvironment for damaged tissues [12, 37]. For treatment, bone marrow aspirate concentration (BMAC) is the major source of MSCs [13, 14], but it has some serious concerns of limitation of volume [13], and inconsistency in progenitor cell counts [38]. To overcome these limitations, *in vitro* culture expansion of MSCs from bone marrow is often used for good clinical effects [39]. However, expanded MSCs in many countries are considered biological drugs and need regulatory approval for their use. Stromal vascular fraction (SVF) or adipose-derived stromal cells (ADSCs) application emerges as a promising alternative to BMAC for non-union fractures[12, 16, 40, 41]. ADSCs/SVF is composed of a heterogeneous population of mesenchymal stem cells (MSCs), fibroblasts, preadipocytes, monocytes, macrophages, lymphocytes, endothelial progenitors, etc., among these MSCs account for 2-10 per cent of ADSCs [42]. Vascular endothelial growth factor, hepatocyte growth factor and other growth factors secreted by adipose derive-MSCs have been shown to promote neovascularization for host tissue repair [43].

Platelets rich plasma (PRP) provide various growth factors and other biomolecules, hence helping in engraftment and growth of administered stem cells. Several reports suggest the augmentative effect of combining PRP and ADSCs for musculoskeletal regeneration increases the proliferation and differentiation of stem cells [44–46]. Few studies have reported that even PRP application alone promotes fracture union; hence, the addition of PRP in our study could potentially confound the results. However, reported results were very mediocre for application of PRP alone in atrophic non-union [47–52]. Despite a heterogeneous group of delayed and non-union and pool having hypertrophic non-unions as well, healing ranged from 30 to 92.86%[47]. When treated with PRP alone, no evidence of union was seen in 12 out of 12 patients (100%) [48], in 6 out of 14 (42.8%) [50] patients and in 7 of 20 cases (35%) [51] despite the types of non-union not being purely atrophic. Another study reported very good outcomes in 14 patients using PRP alone, but all these non-unions were oligotrophic [49]. Another study, which showed promising results by using PRP alone, reported union in 82 out of 94 patients (87%) within 4 months; however, the patient group was heterogeneous and lacked pure atrophic cases [52]. Therefore, we choose to combine PRP in ADSC to aid stem cell growth and proliferation, since we understand that PRP alone is not therapeutic in atrophic nonunion.

The non-enzymatic ultrasonic cavitation method of ADSCs isolation [29] used in this study is considered to exhibit minimal manipulation and is hence compliant with regulatory laws. The isolation process yielded 14.57-17.14 million stromal cells in one mL of the specimen, which implies a good yield of 116-137 million cells from 100 mL of lipoaspirate.

Ultrasonication causes cavitation, which disrupts the extracellular matrix and homogenizes adipose tissue. Adipose cells are more sensitive to sonication-mediated cavitation than stromal cells, making them easier to disrupt [29, 53]. Similar to collagenase, sonication is highly efficient in releasing collagen fibers that tightly anchor fat tissue cells [54]. Although ultrasound-based techniques for stromal vascular fraction (SVF) isolation have been proposed, they are not yet widely adopted in clinical practice [55, 56].

Existing SVF isolation methods using ultrasonic cavitation report yields ranging from 1.67 to 2.24 × 10LJ cells, with viability rates of 97.1–98.9% [57]. Bright et al. report yields of 2–4 million cells per gram of adipose tissue[58]. Gao et al. achieved yields of 2.70 × 10LJ cells/mL [59], similar to the enzymatic methods reported by Park et al [60]. Sonication methods have demonstrated nucleated cell yields from 7 × 10LJ to 10 × 10LJ cells per gram of adipose tissue [61], and 2.6 × 10LJ viable nucleated cells/mL of lipoaspirate—slightly higher than the 2.0 × 10LJ total nucleated cells/mL obtained using enzymatic methods [29, 62].

Ultrasonic cavitation also results in a greater number of exosome particles [62]and a higher proportion of ADSCs—more than 10%—compared to other mechanical approaches that yield only 1%–5% ADSCs in the SVF composition [29, 60]. There is generally no significant difference in the expression of stem cell CD markers between enzymatic and ultrasound methods [60, 62, 63]. However, the expression of positive markers such as CD34, CD73, and CD105 is more robust in ADSCs isolated via sonication, with relatively lower expression of hematopoietic markers like CD45 and CD11b and a higher expression of CD105 compared to enzymatic processes [60, 63].

Functionally, ADSCs isolated using ultrasonication exhibit similar adipogenic, osteogenic, and chondrogenic differentiation potential to those isolated enzymatically [29, 59, 60]. Growth trends, including proliferation rates and colony-forming unit-fibroblast (CFU-F) formation, were also comparable between the collagenase and ultrasound methods, with only minimal differences reported [59, 60].

Furthermore, clinical applications using AD-SVFs obtained via ultrasonic cavitation have demonstrated safety and efficacy. Systemic infusion in patients with chronic migraine and intra-articular administration of 50 to 100 million cells in osteoarthritis (OA) patients were found to be safe and therapeutically effective [61, 64].

Patient-derived ADSCs were freshly isolated and characterised by flow cytometry to evaluate their stemness and multilineage differentiation capacity based on the expression of specific positive and negative surface markers. The results show positive expression of classical ADSC markers CD29, CD90, CD105, and CD73, consistent with earlier reports [31, 65–67].

After injecting autologous ADSCs suspended in PRP into the patient’s fracture site, six of the seven patients achieved union, and five showed the first evidence of healing by eight weeks after the procedure. The clinical improvement corresponded well with the X-ray imaging. The most difficult situation was with patient P7, who had a gap of almost 5mm. Hence, the time taken by this patient to heal was longer when compared to others.

Our results corresponded well with previous studies using bone marrow concentrate, which is still the most frequently used cell-based bone regeneration procedure. Studies have reported union in 78% to 100% of patients in closed [10, 14] and 53.5% in open tibial non-unions [13]. Despite a larger fracture gap in our study, the median time to union was 10.6 weeks, comparable to the healing timelines reported in studies where bone marrow concentrate was used [10, 13, 14, 68, 69]. It is noteworthy that our selected patients had a duration of non-union ranging from 9-16 months. It can be asserted that union in these cases would not have occurred without this procedure, as all the patients had an occluded marrow cavity with sclerosis at the bone ends, and the mean duration between the ADSC procedure and the duration since injury was 11.7 months. No major local or systemic complications were observed. One patient experienced short-lived local pain and swelling at the liposuction site. We performed implantation of ADSCs in patients without removing the fibrous tissue interposed between the bone ends was ossified. The exact mechanism of the transformation of fibrous tissue into callus remains unclear. Although the intrinsic potential of a local tissue is generally impaired at the atrophic non-union site, its regenerative cells can be re-stimulated to form bone structures under appropriate conditions. This phenomenon suggests that progenitor cells in atrophic non-union tissues can regenerate bone even though the microenvironment in the non-union site might retard their function. Some physical or biological agents (like growth factors or cells) are required to reactivate the endogenous progenitor cells to revive them towards bone regeneration in atrophic non-union [22]. The “cocktail” of preadipocytes, pericytes, MSCs, and various endothelial progenitor cells present in SVF [70] provides angiogenic, anti-inflammatory, and immune modulation effects. MSCs residing in the SVF possess strong osteogenic potential not only by their ability to differentiate into osteogenic cells following induction but also by their paracrine effects through the secretion of numerous growth factors and cytokines [39, 71]. These factors influence the encompassing progenitor cells. Some evidence-based studies suggest that these infused stem cells get engrafted into the tissue and differentiate into tissue [72].

The present study is a proof-of-concept, the first ever detailed report on the efficacy of ADSCs in the treatment of long bone non-union. The first successful case with ADSCs implantation was reported for extensive calvarial injury in a seven-year-old girl [22], followed by a few reports of success in treating skull and jaw defects [20, 73, 74]. Consolidation in 80% and 50% of patients with post sarcoma long bone defects was reported using ADSCs and bone matrix [23, 24]. The first ever non-union treated with ADSCs was a sternal restoration, which showed healing by the end of 3 months on 3D-CT imaging [75]. A positive outcome of SVF implantation in 8 cases with fresh proximal humeral fractures in severe osteoporotic bones has also been reported.

The major limitation of our study was the small sample size to reach any definitive conclusions, despite the study assessments having a long follow-up. The study was also limited as all the patients received almost the same dose (approx. 100 million cells) irrespective of age, weight, duration of non-union, the width of bone and fracture type. Validation with more patients would bring more conclusive evidence. However, we can say beyond doubt that the ADSCs application is a possible alternative tool for managing non-union.

## Conclusion

Our results strongly suggest that ADSCs administration is a safe, effective, and promising therapeutic tool and a preferable alternative to bone marrow concentrate in treating non-union fractures of long bones. Clinical results indicate the application of ADSCs can also be extended to delayed union or in patients where chances of nonunion are high during primary fixation. However, more clinical studies with a greater number of subjects are warranted to study the comparative efficacy and other practical challenges of the treatment.

## List of abbreviations

ADSC: Adipose derived stromal cell
BMAC: Bone Marrow Aspirate Concentrate
BM-MSC: Bone Marrow derived Mesenchymal Stem Cells
CFU-F: Colony-forming-unit fibroblast
IM: Intramedullary
LEFS: Lower Extremity Functional Scale
MSC: Mesenchymal Stem Cells
POP: Plaster of pairs
PRP: Platelet Rich Plasma
SF-12: Short Form 12
SVF: Stromal Vascular Fraction

## Declarations

### Institutional Review Board Statement

This trial was ethically approved by the Institutional Committee for Stem Cell Research and Therapy, Anupam Hospital, Uttarakhand, India. The trial is compliant with the consolidated standards of reporting trials (CONSORT). Informed prior consent was obtained from all the patients. This trial was registered in Clinicaltrials.gov-NCT04340284; Date of registration 09/04/2020; Clinicaltrials.gov under URL: https://clinicaltrials.gov/ct2/show/NCT04340284).

## Contributions

H.B. has designed and conducted the study. A.B drafted the study protocol; A.B and A.B. assisted in designing the protocol, supervising the study and reviewing the results. A.B. supervised the entire study and treated the selected patient population. H.B and I.K writing— review and editing, K.C and J.L. has reviewed the X-ray and MRI of patients. SS has helped in fund acquisition and Project administration. All authors have read and agreed to the published version of the manuscript.

## Informed Consent Statement

Informed consent was obtained from all subjects involved in the study. Written informed consent has been obtained from the patients to publish this paper.

## Data Availability Statement

The data presented in this study are available on request from the corresponding author.

## Data Availability

All data produced in the present study are available upon reasonable request to the authors

## Acknowledgements

We are thankful to the patients for their kind collaboration. The authors are thankful to the laboratory staff for collecting and biobanking patients’ samples.

